# Acute Ischemic Stroke in Tsutsugamushi: Understanding the Underlying Mechanisms and Risk Factors

**DOI:** 10.1101/2023.06.06.23291000

**Authors:** Dain Kim, Yeon Hee Cho, Ji Yeon Chung, Young Seo Kim, Byoung-Soo Shin, Hyun Goo Kang

## Abstract

**Background:** Tsutsugamushi, also known as scrub typhus, is an acute infectious febrile disease common in the Asia–Pacific region. Its common symptoms include lymphadenopathy, fever, and myalgia. Tsutsugamushi rarely causes acute ischemic stroke (AIS); however, it has been reported to increase the likelihood of AIS development. Here, we aimed to test the hypothesis that tsutsugamushi infection could trigger AIS.

**Method:** We retrospectively examined patients diagnosed with AIS within 2 weeks of tsutsugamushi diagnosis at three hospitals over 15 years. We categorized patients who developed AIS while being treated for tsutsugamushi as the case group; patients who did not develop AIS while being treated for tsutsugamushi at the same hospital and were of similar age and sex with the case group were categorized as the control group. The case and control groups had 22 and 66 participants, respectively. When a scattered pattern was observed or lesions were found in two or more vascular territories on diffusion-weighted imaging, the pattern was defined as embolic. Other patterns were defined as non-embolic.

**Results:** Among the 19 patients, excluding three with transient ischemic attack, 15 (78.9%) showed an embolic pattern. Fever was common at the time of the visit in the control group; however, it was less common in the case group (93.8% vs. 27.3%, respectively; p<0.001). Multivariate analysis revealed that higher D-dimer levels at the time of hospitalization were associated with AIS development in patients with tsutsugamushi (adjusted odds ratio, 1.58; 95% confidence interval, 1.06–2.34; p=0.023).

**Discussion:** AIS in patients with tsutsugamushi showed an embolic rather than a non-embolic pattern on brain magnetic resonance imaging. This embolic pattern was more likely to occur in patients with stroke risk factors. These patients were unlikely to have a fever or high D-dimer levels. We speculated that D-dimers played a vital role in the pathophysiology, as tsutsugamushi infection increased the likelihood of AIS.

## Introduction

Tsutsugamushi, also known as scrub typhus, is an acute infectious febrile disease caused by the bacterium *Orientia tsutsugamushi*. The disease is prevalent in Asia and the Pacific and Indian Ocean islands.^1^ Particularly, in 2022, 6,232 patients with tsutsugamushi were reported in South Korea, with approximately 5,000 cases per year in the last 5 years, thus showing its high prevalence.^2^

Common symptoms of tsutsugamushi include eschar, lymphadenopathy, fever, headache, myalgia, and gastrointestinal symptoms, whereas severe symptoms include interstitial pneumonia, acute respiratory distress syndrome, meningoencephalitis, acute kidney injury, and disseminated intravascular coagulation.^1^ Acute ischemic stroke (AIS) is a rare symptom of tsutsugamushi; however, some patients present with both tsutsugamushi and AIS in clinical practice. Such cases have been reported infrequently. We hypothesized that tsutsugamushi infection could be a causal factor of AIS because previous studies have reported that acute infections, like tsutsugamushi, could increase the occurrence of AIS.^3^ This study aimed to investigate the specific mechanisms underlying the development of AIS in patients with tsutsugamushi.

The primary pathophysiology of tsutsugamushi infection affecting blood vessels involves endothelial damage and vasculitis. However, given that AIS rarely develops in patients with tsutsugamushi, we surmised there might be other mechanisms besides vasculitis. We hypothesized that the coagulation disorder induced by tsutsugamushi infection leads to AIS, based on the results obtained from previous studies, which suggested that tsutsugamushi could cause a coagulation pathway disorder^4^ and that thrombogenesis is the infection’s main mechanism of inducing stroke.^5^ To test this hypothesis, we analyzed the data of tsutsugamushi patients with AIS.

## Methods

### Patient selection

This study retrospectively evaluated patients hospitalized for tsutsugamushi at three regional hub hospitals in different urban areas over 15 years (2005–2019) and who were diagnosed with AIS within 2 weeks of tsutsugamushi diagnosis. We collated and analyzed patients’ symptoms, medical history, and laboratory findings at admission using the electronic medical record. We categorized patients who developed AIS while being hospitalized and treated for tsutsugamushi as the case group. Those patients who did not develop AIS while being hospitalized and treated for tsutsugamushi at the same hospital and were similar in age and sex were categorized as the control group. As a result, this study categorized 22 and 66 patients into the case and control groups, respectively. This study was conducted following the Declaration of Helsinki and approved by the Institutional Review Board at Jeonbuk National University Hospital (CUH 2023-05-001). The requirement for informed consent was waived owing to the retrospective nature of the study.

### Definition

Patients were diagnosed with tsutsugamushi when the *O. tsutsugamushi* gene was detected using polymerase chain reaction, *O. tsutsugamushi* was identified in the serum, or IgM antibodies against *O. tsutsugamushi* were detected. The AIS group included patients with AIS confirmed by brain magnetic resonance imaging (MRI) and patients with transient ischemic stroke (TIA). However, this study excluded patients with subacute or chronic stroke. TIA was defined as stroke-related neurological symptoms, such as dysarthria or hemiplegia, which improved within 24 h. Patient symptoms were identified based on the presenting symptoms at hospitalization. This study examined basic demographic information (age and sex), the presence of eschar owing to tsutsugamushi, and various tsutsugamushi-related symptoms (fever, headache, fatigue, anorexia, dyspepsia, nausea, vomiting, abdominal pain, diarrhea, arthralgia, myalgia, sore throat, back pain, dyspnea, cough, and thirst). In addition, we recorded the antibiotics used to treat tsutsugamushi, measured the time from admission to the development of AIS symptoms, and investigated the risk factors for stroke (hypertension, diabetes mellitus, atrial fibrillation, dyslipidemia, coronary arterial disease, drinking history, and smoking history) at admission. Regarding stroke risk factors, a patient was defined to have a risk factor as an underlying disease if the patient had been diagnosed with a risk factor and received medication for it or met the relevant diagnostic criteria after hospitalization, even though the patient had no risk factor diagnosis. Hypertension was defined as when the blood pressure measured at rest during hospitalization was _130/80 mmHg.^6^ Diabetes mellitus was diagnosed if the level of 8-h fasting blood glucose was ≥126 mg/dL, HbA1c was ≥6.5%, or blood glucose was ≥200 mg/dL after 2 h of oral glucose tolerance test.^7^ Atrial fibrillation included all patients who had atrial fibrillation confirmed by electrocardiogram performed after admission or those previously diagnosed with atrial fibrillation. Dyslipidemia was diagnosed if the level of low-density lipoprotein was ≥160 mg/dL, triglyceride was ≥200 mg/dL, high-density lipoprotein was ≤40 mg/dL, or total cholesterol was ≥240 mg/dL.^8–10^ Coronary artery disease was diagnosed if coronary angiography confirmed abnormal findings. If a patient’s response showed that the patient had consumed alcohol or cigarettes within 1 year of hospitalization, the patient was defined as having a drinking or smoking risk factor. Aspartate aminotransferase (AST), alanine aminotransferase (ALT), white blood cell (WBC), hemoglobin (Hg), and platelet (PLT), which are laboratory findings generally known to show abnormal values in patients with tsutsugamushi,^11^ and prothrombin time (PT), activated partial thromboplastin time (aPTT), D-dimer, fibrinogen, and fibrin degradation product (FDP), which are laboratory findings related to blood coagulation that may affect ischemic stroke, were investigated based on blood tests at hospitalization.

### Imaging analysis

This study investigated the pattern of lesions caused by stroke by collecting and analyzing brain MRIs of patients admitted for tsutsugamushi and diagnosed with AIS or TIA during treatment. Subsequently, we examined the presence of atrial fibrillation and D-dimer levels, which are factors believed to be associated with embolic patterns. When a scattered pattern (indicating that a high signal intensity is scattered over multiple territories) was observed on diffusion-weighted imaging (DWI) or when there were lesions in two or more different vascular territories, it was defined as an embolic pattern. Other types were defined as non-embolic.

### Statistical analysis

First, a control group was selected based on the age and sex matching of patients with a disposition similar to that of the case group. Age and sex matching were performed using propensity score matching and the nearest neighbor matching algorithm, and 66 patients were selected as the control group using a 1:3 matching ratio. Afterward, we compared the demographics, clinical symptoms, and stroke risk factors of patients with tsutsugamushi with or without AIS. We used Pearson’s chi-squared or Fisher’s exact tests to analyze categorical variables, and Student’s t-test was used for continuous variables. Continuous variables were summarized as means with standard deviations or medians with interquartile ranges, whereas categorical data were expressed as counts and percentages. Multivariate analysis was performed to identify the independent factors associated with tsutsugamushi presenting with AIS. Only variables showing a potential association (p<0.1) in univariate analysis were included as potential factors associated with tsutsugamushi presenting with AIS in the multivariate logistic regression model to avoid variable selection caused by spurious correlations. Statistical significance was set at p<0.05 (two-tailed). All statistical analyses were performed using SPSS 21.0 (IBM Corporation, Armonk, NY, USA).

## Results

Among the 22 patients with tsutsugamushi who developed AIS, 14 were women (63.6%), and 8 were men (36.4%); the mean age was 74.4 ± 13.4 years, with the youngest patient being 26 years old and the oldest being 93 years old (Table 1). Antibiotics used for treating tsutsugamushi included doxycycline in 16 patients (72.7%), azithromycin in four (18.2%), and rifampicin in two (9.1%). The average time from the onset of tsutsugamushi symptoms to that of AIS was 3.91 ± 4.54 days; however, the onset time in four patients was _10 days. In addition, factors associated with vascular risk were current alcohol consumption in two (9.1%) participants, current smoking status in two (9.1%), hypertension in 12 (54.5%), dyslipidemia in seven (31.8%), diabetes mellitus in five (22.7%), atrial fibrillation in five (22.7%), and coronary artery disease in two (9.1%).

**Table 1.**
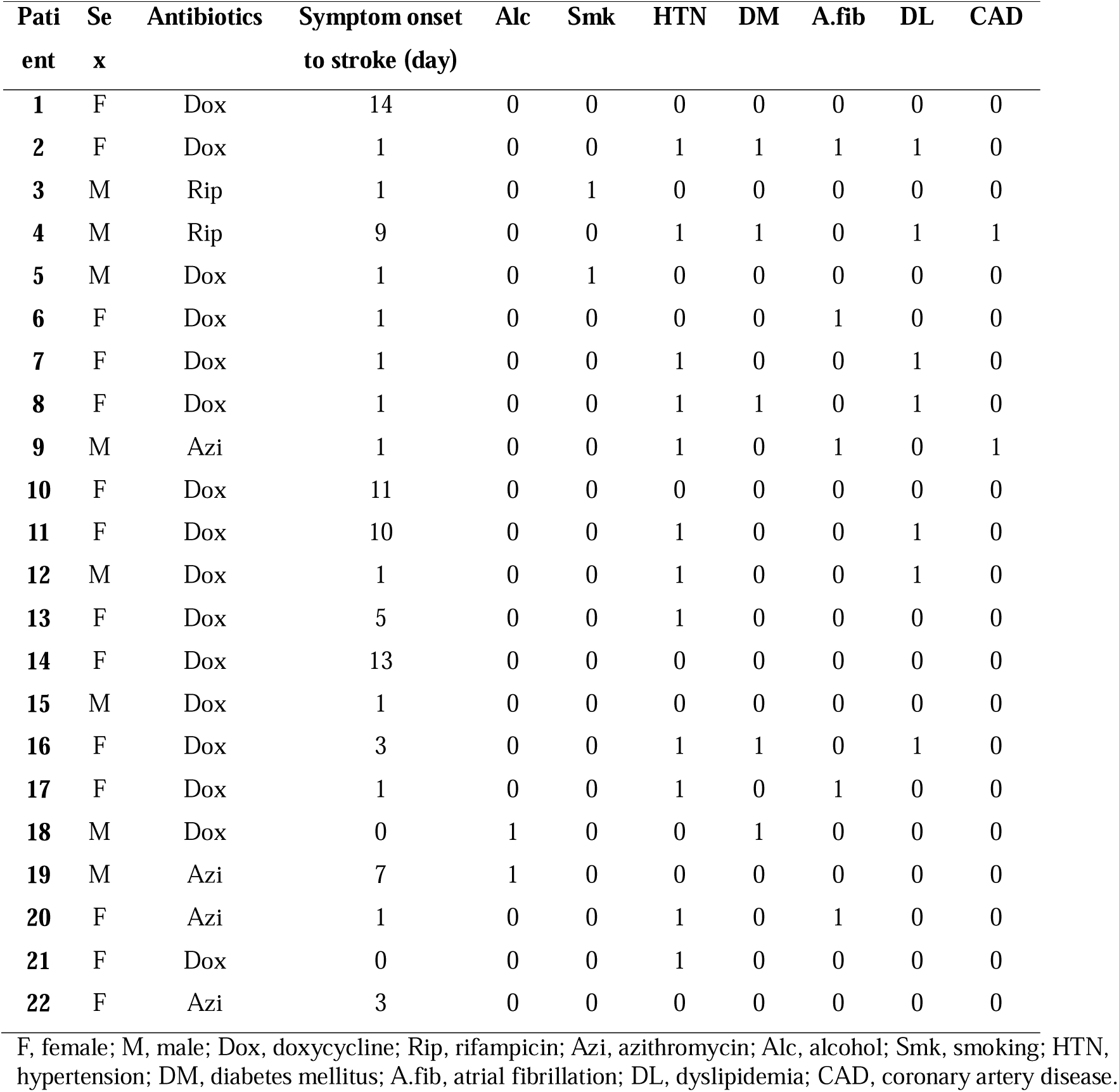
Baseline characteristics of patients with tsutsugamushi and acute ischemic stroke

This study described the brain MRI characteristics of 19 patients with tsutsugamushi and AIS, excluding three patients with TIA with no lesions on DWI (Table 2). Among the 19 patients, 15 (78.9%) showed an embolic pattern, whereas four (21.1%) had a non-embolic pattern, indicating that an embolic pattern was more common. Sixteen (84.2%) patients had elevated D-dimer levels above the normal level (0.5 mg/L), and six (31.6%) had atrial fibrillation as an underlying condition. Among the 16 patients with elevated D-dimer levels, 13 (81.3%) had an embolic pattern, and five (31.3%) had atrial fibrillation. Five of the six patients with atrial fibrillation had elevated D-dimer levels, and one did not; all six patients showed an embolic pattern. None of the 19 patients had a history of cancer.

**Table 2.**
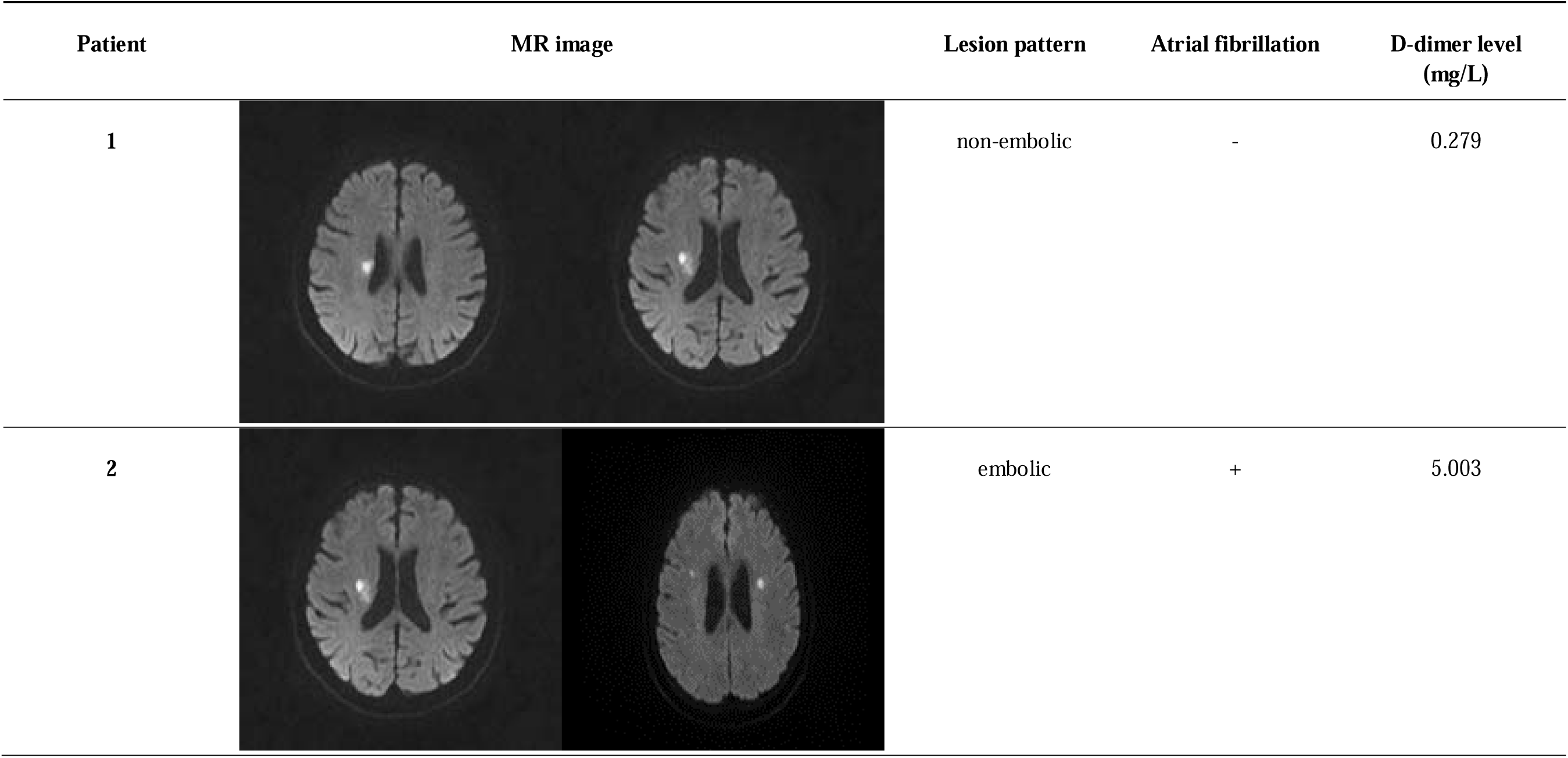
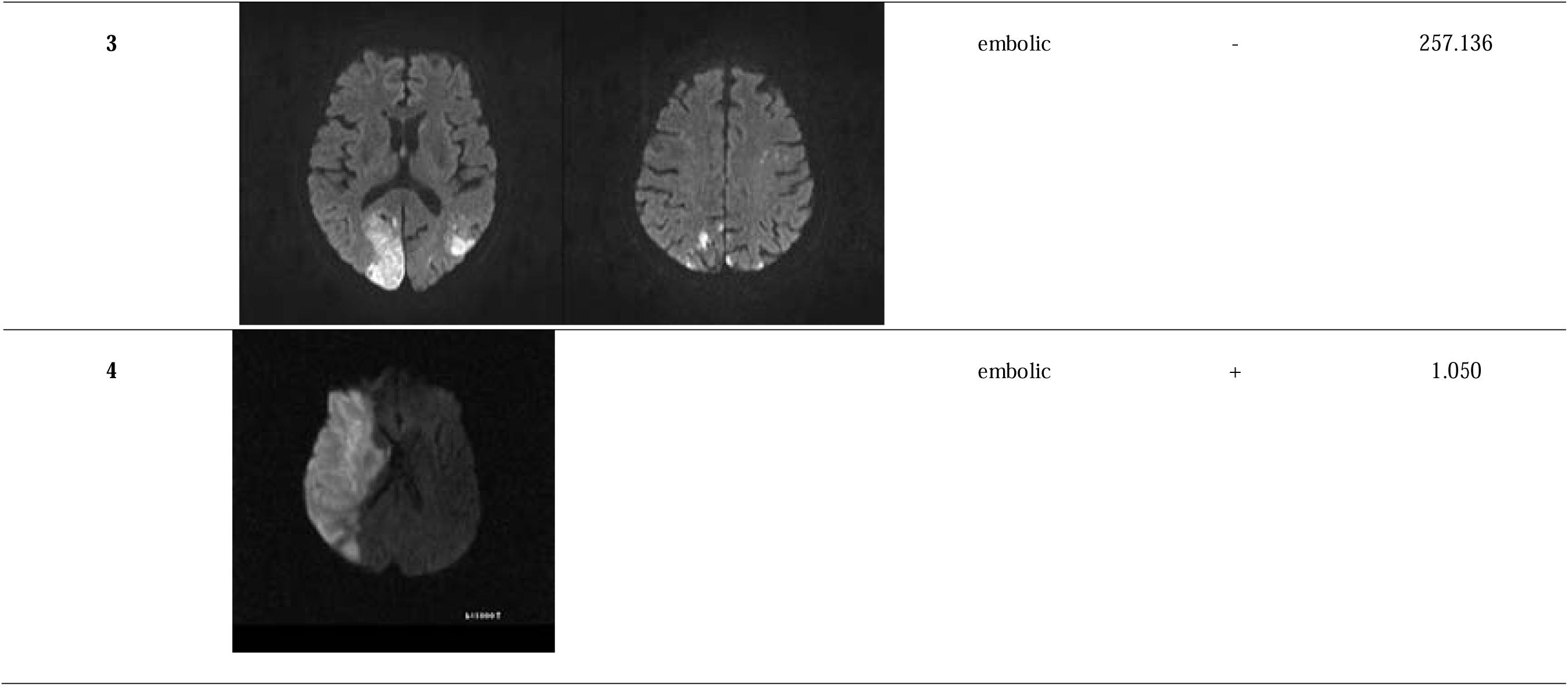
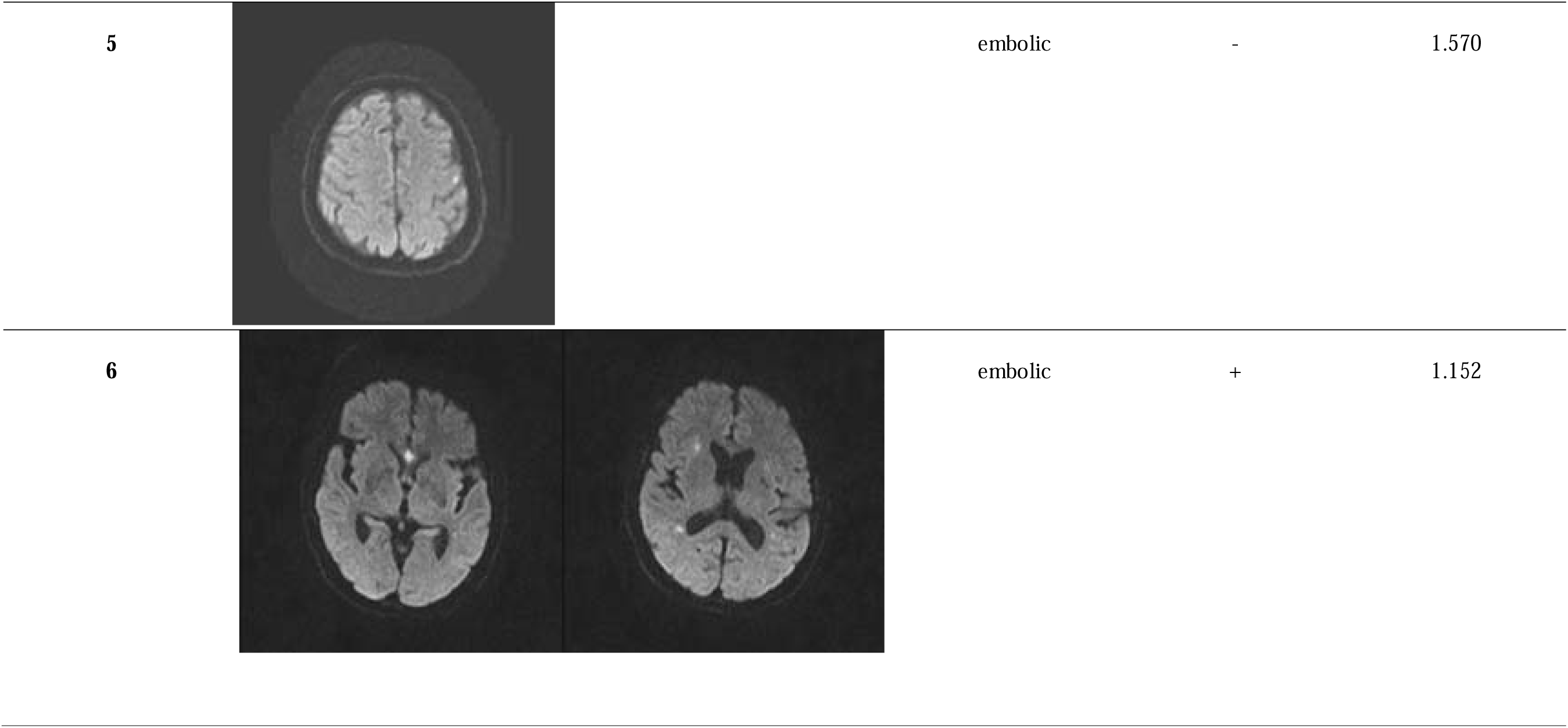
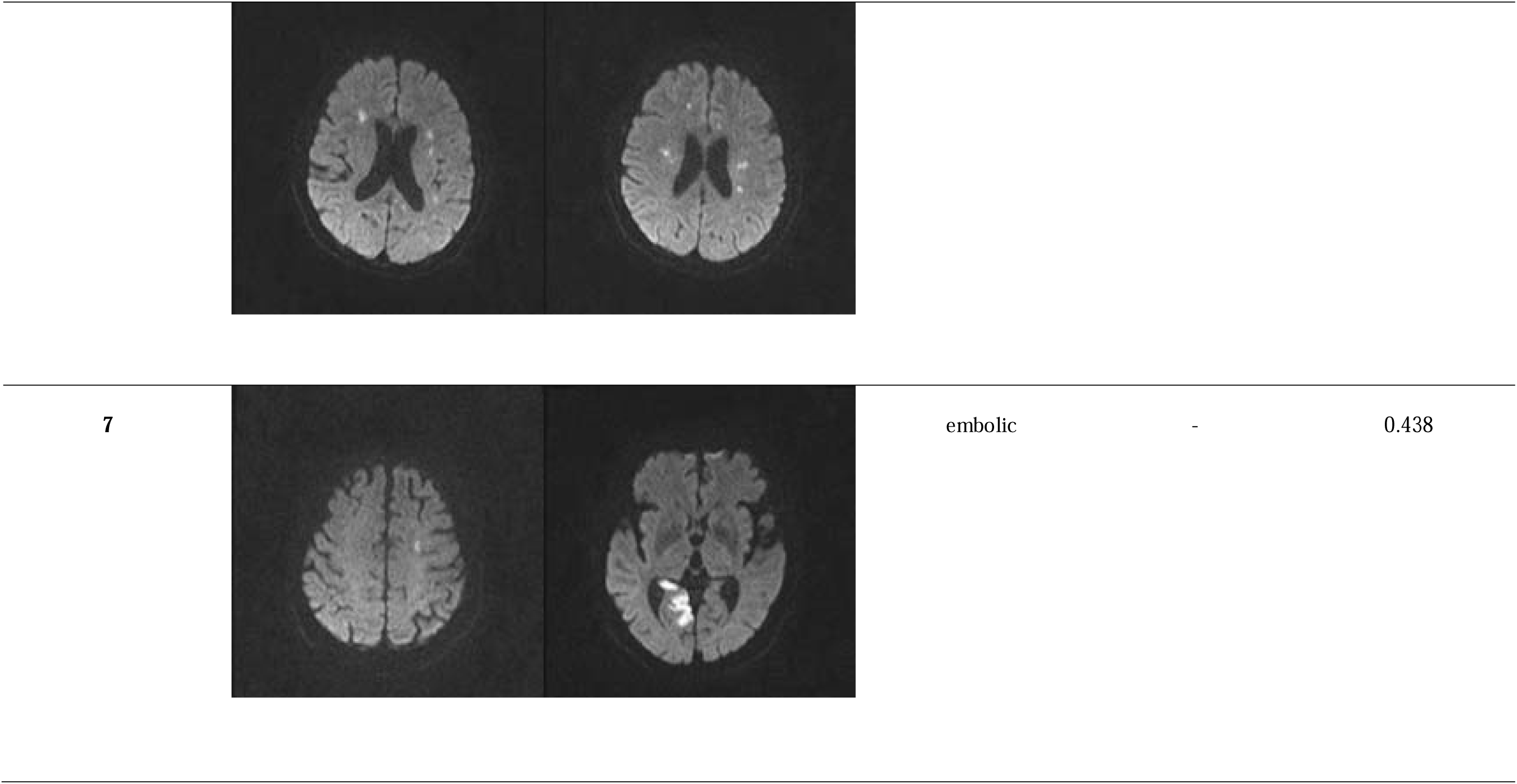
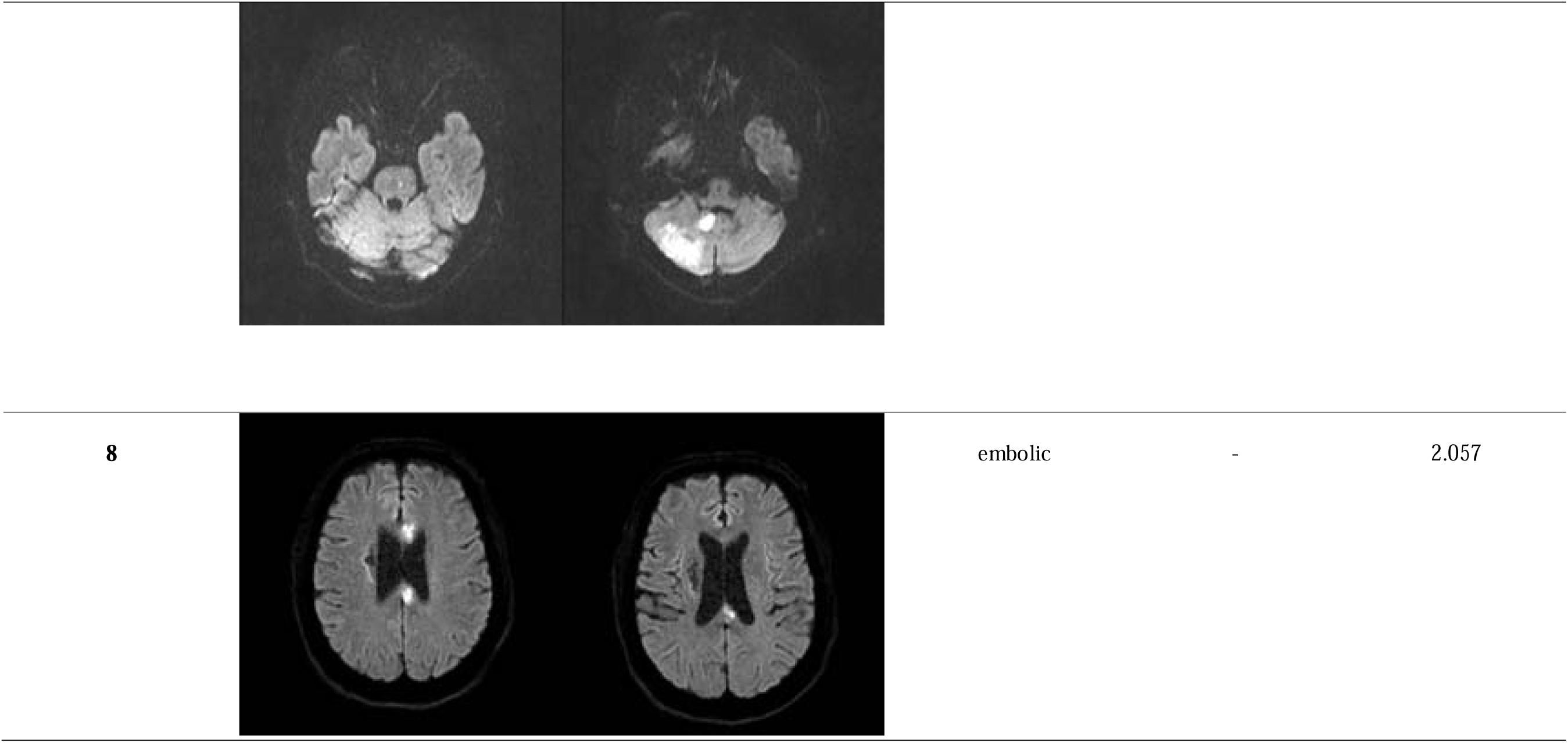
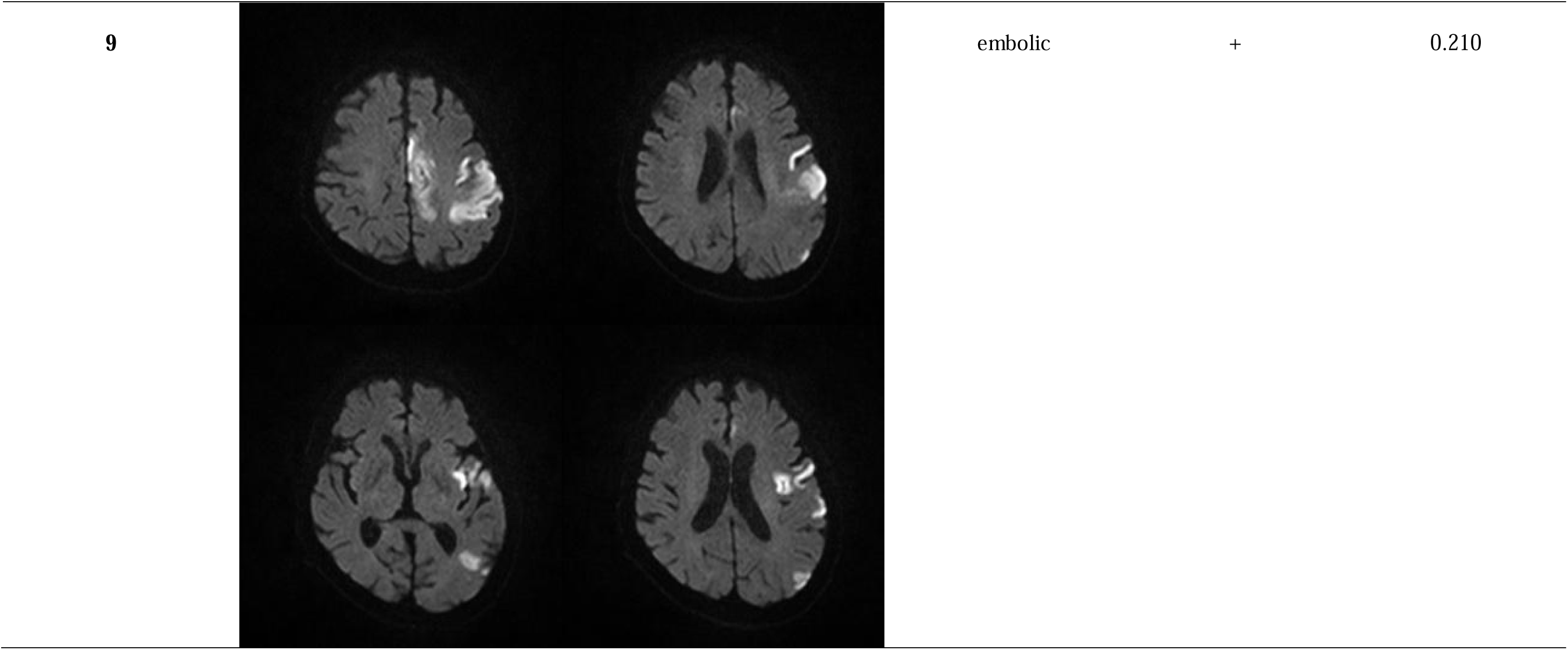
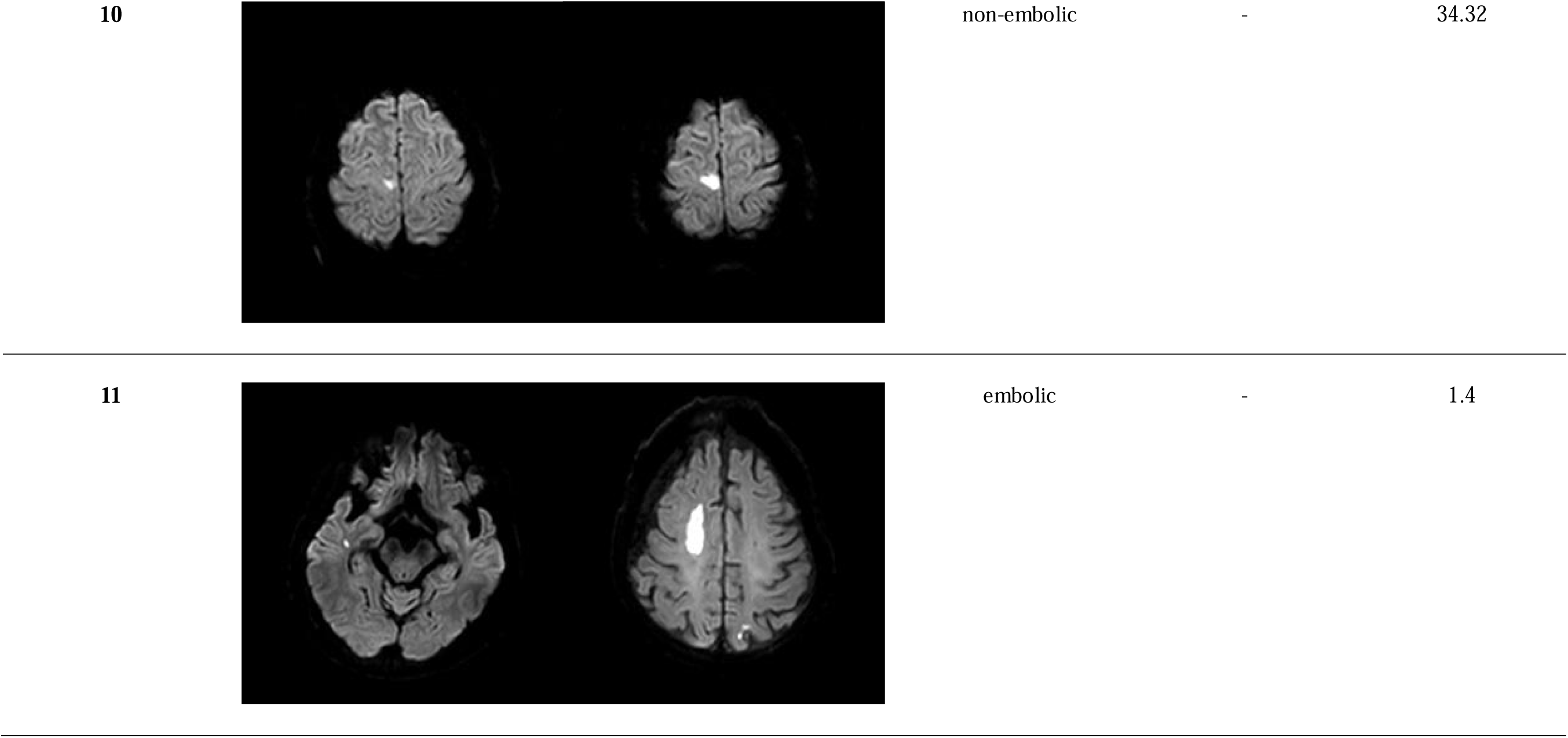
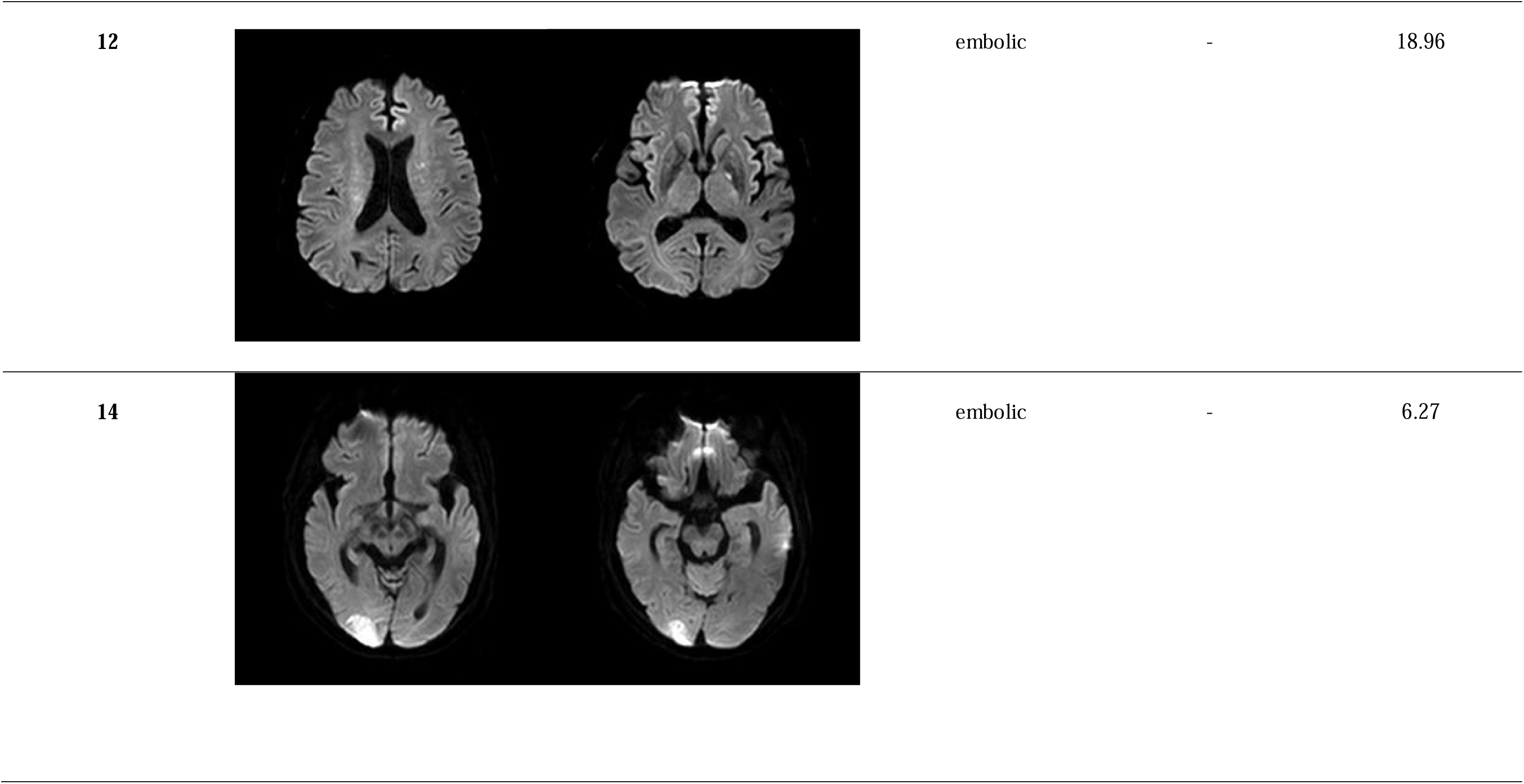
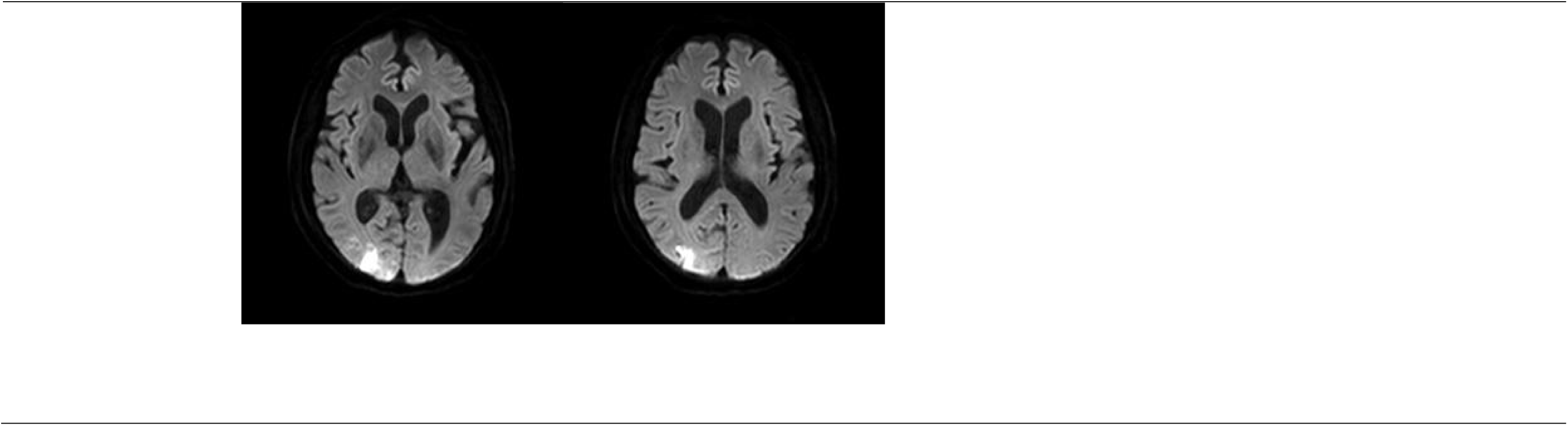
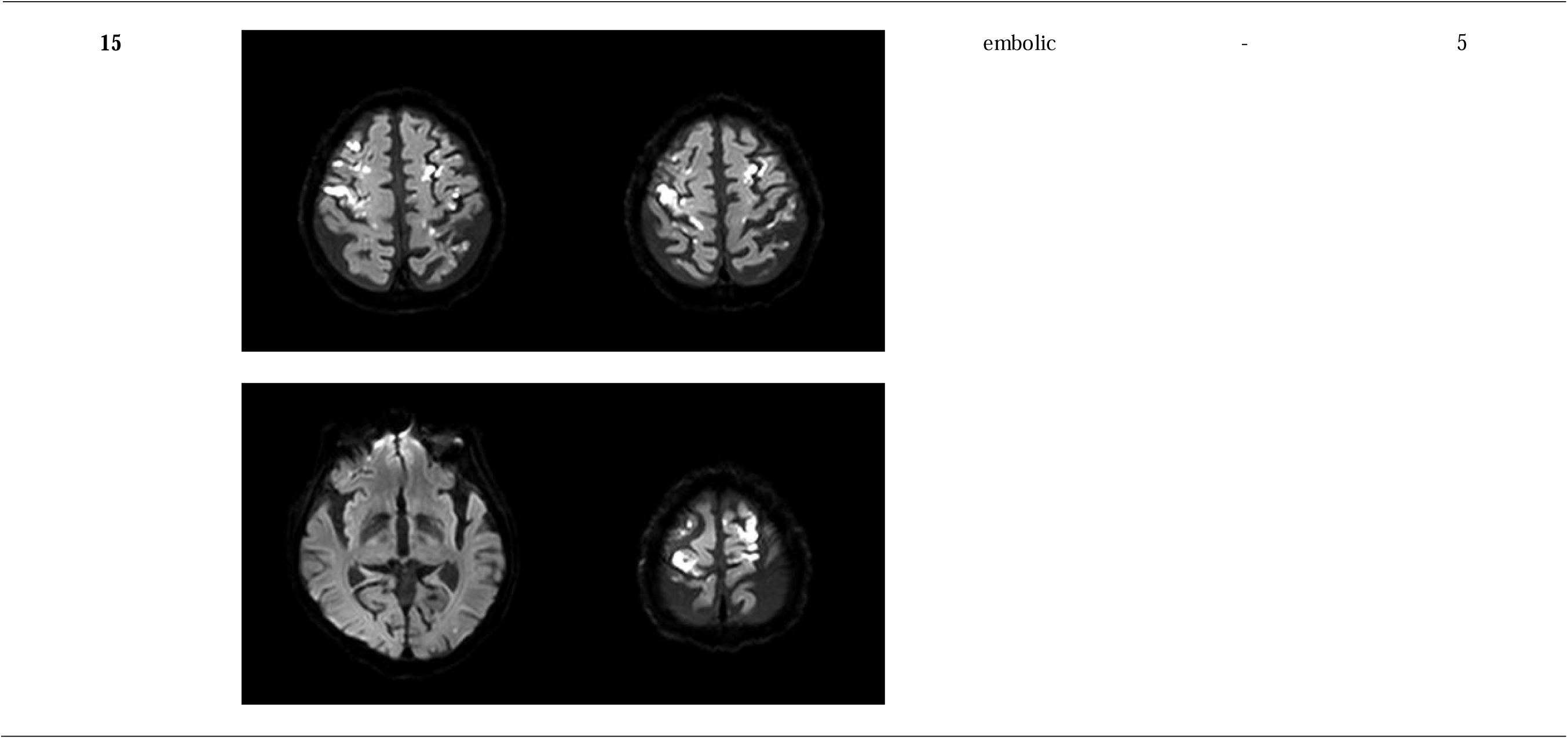
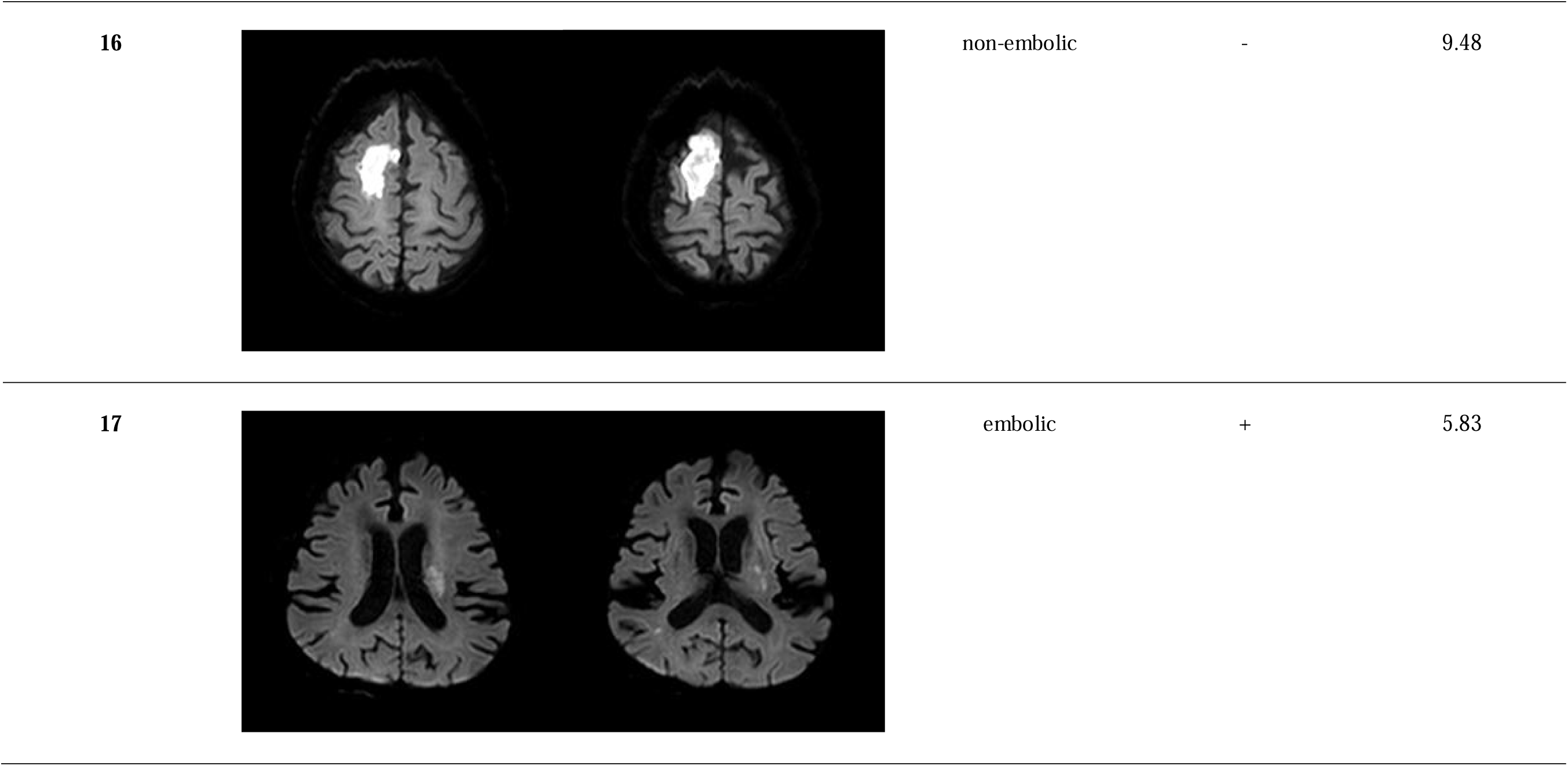
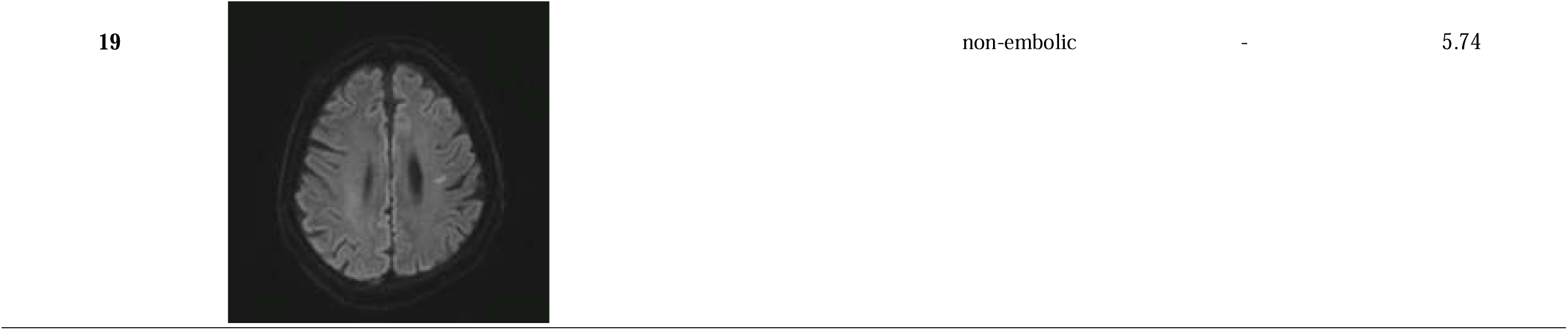
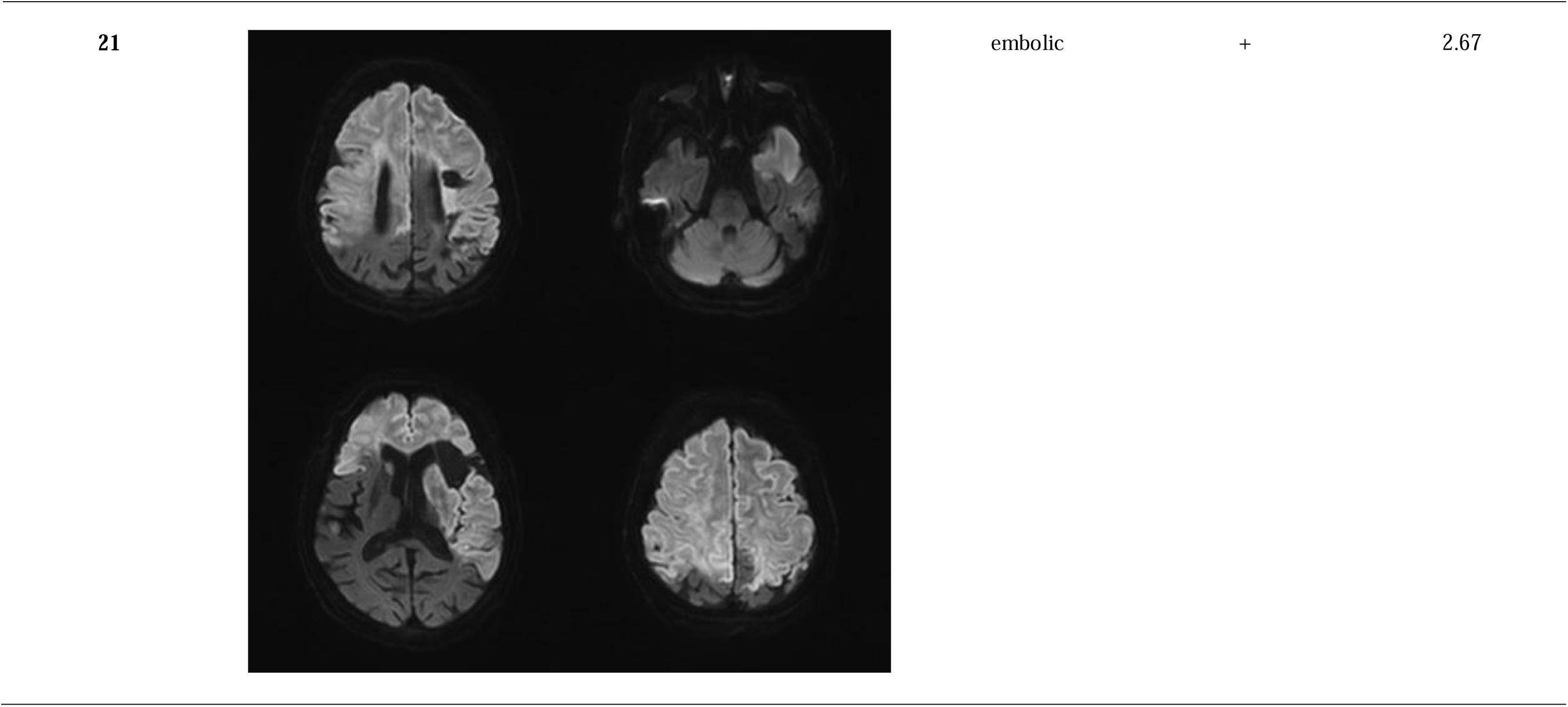
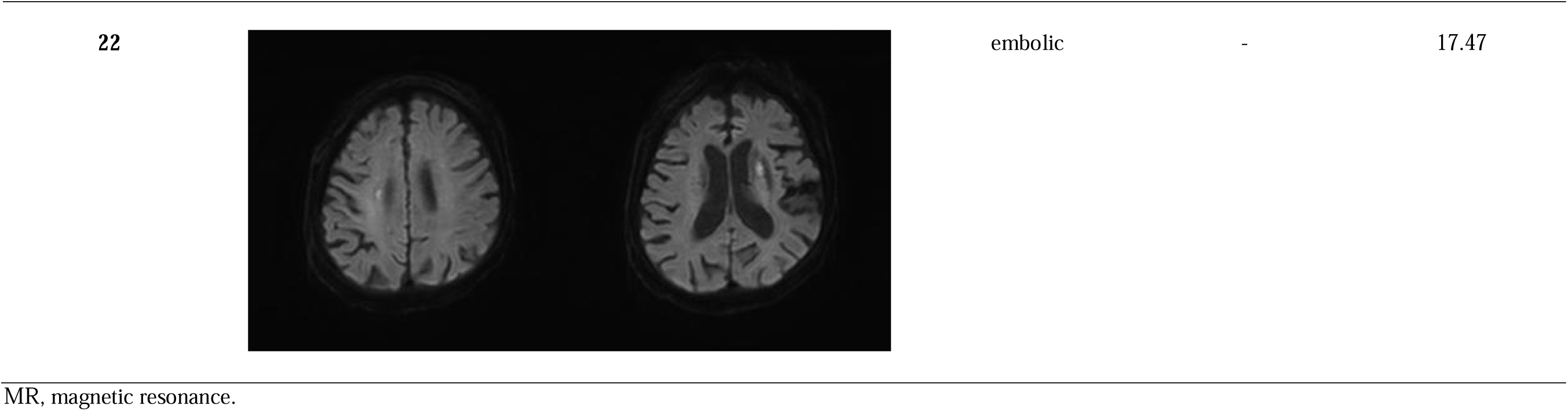
Diffusion-weighted imaging of a patient with acute ischemic stroke, with tsutsugamushi and stroke patterns (embolic, non-embolic) and risk factors of the patients

This study compared patients with tsutsugamushi with and without AIS (Table 3). The mean age of the target group was 74.41 ± 13.36 years, compared with 74.18 ± 12.81 years of the control group. At admission, fever was commonly observed in patients with tsutsugamushi without AIS but was significantly less common in tsutsugamushi patients with AIS (93.8 vs. 27.3%, respectively; p<0.001). In addition, headache was significantly less common in tsutsugamushi patients with AIS than in those without AIS (tsutsugamushi patients without AIS vs. tsutsugamushi patients with AIS; 69.8 vs. 31.8%, respectively; p=0.002). The incidence of sore throat (tsutsugamushi patients without AIS vs. tsutsugamushi patients with AIS; 31.8 vs. 9.1%, respectively; p=0.036), dyspnea (21.2 vs. 0%, respectively; p=0.018), and thirst (82.9 vs. 25.0%, respectively; p<0.001) was lower in tsutsugamushi patients with AIS than in those without AIS. Tsutsugamushi patients with AIS were more likely to have stroke risk factors than those without AIS. Notably, tsutsugamushi patients with AIS were significantly more likely to have hypertension (tsutsugamushi patients without AIS vs. tsutsugamushi patients with AIS; 16.1 vs. 54.5%, respectively; p=0.001), diabetes mellitus (5.3 vs. 22.7%, respectively; p=0.021), atrial fibrillation (4.5 vs. 22.7%, respectively; p=0.01), and dyslipidemia (7.6% vs. 31.8%, respectively; p=0.004) than did tsutsugamushi patients without AIS. Regarding laboratory findings, tsutsugamushi patients with AIS showed significantly higher levels of aPTT (tsutsugamushi patients without AIS vs. tsutsugamushi patients with AIS; 32.31 ± 12.27 vs. 40.54 ± 13.47, respectively; p=0.009), D-dimer (0.87 ± 1.27 vs. 8.14 ± 10.47, respectively; p=0.005), and FDP (2.27 ± 0.69 vs. 42.01 ± 59.55, respectively; p=0.009) than did tsutsugamushi patients without AIS. The results in Table 3 are presented as a bar graph (Figure 1).

**Table 3.** Comparison of tsutsugamushi with and without acute ischemic stroke

|  | Tsutsugamushi without stroke<br>(n=66) | Tsutsugamushi with stroke<br>(n=22) | *P value* |
| --- | --- | --- | --- |
| **Female** | 39 (59.1) | 14 (63.6) | *0.706* |
| **Age** | 74.18 ± 12.81 | 74.41 ± 13.36 | *0.943* |
| **Eschar** | 52 (83.9) | 17 (77.3) | *0.524* |
| **Symptoms** |  |  |  |
| Fever | 61 (93.8) | 6 (27.3) | *<0.001* |
| Headache | 44 (69.8) | 7 (31.8) | *0.002* |
| Fatigue | 39 (59.1) | 17 (77.3) | *0.125* |
| Anorexia | 46 (69.7) | 13 (59.1) | *0.359* |
| Dyspepsia | 16 (41.0) | 7 (33.3) | *0.559* |
| Nausea | 15 (29.4) | 4 (18.2) | *0.316* |
| Vomiting | 6 (11.8) | 3 (13.6) | *1* |
| Abdominal pain | 5 (7.6) | 1 (4.5) | *1* |
| Diarrhea | 2 (3.0) | 2 (9.1) | *0.259* |
| Arthralgia | 14 (22.2) | 5 (23.8) | *1* |
| Myalgia | 38 (60.3) | 12 (54.5) | *0.636* |
| Sore throat | 21 (31.8) | 2 (9.1) | *0.036* |
| Back pain | 7 (10.6) | 2 (9.1) | *1* |
| Dyspnea | 14 (21.2) | 0 (0.0) | *0.018* |
| Cough | 15 (23.8) | 2 (10.5) | *0.335* |
| Thirst | 29 (82.9) | 4 (25.0) | *<0.001* |
| **Stroke risk factors** |  |  |  |
| Hypertension | 9 (16.1) | 12 (54.5) | *0.001* |
| Diabetes mellitus | 3 (5.3) | 5 (22.7) | *0.021* |
| Atrial fibrillation | 3 (4.5) | 5 (22.7) | *0.01* |
| Dyslipidemia | 5 (7.6) | 7 (31.8) | *0.004* |
| Coronary artery disease | 3 (5.5) | 2 (9.1) | *0.612* |
| Alcohol | 2 (5.1) | 2 (9.1) | *0.615* |
| Smoking | 3 (7.9) | 2 (9.1) | *1* |
| **Laboratory findings** |  |  |  |
| PT | 12.47 ± 1.06 | 14.18 ± 7.53 | *0.302* |
| aPTT | 32.31 ± 12.27 | 40.54 ± 13.47 | *0.009* |
| D-dimer | 0.87 ± 1.27 | 8.14 ± 10.47 | *0.005* |
| Fibrinogen | 292.96 ± 98.15 | 252.62 ± 106.69 | 0.132 |
| FDP | 2.27 ± 0.69 | 42.01 ± 59.55 | 0.009 |
| AST | 194.18 ± 860.00 | 99.82 ± 113.20 | 0.611 |
| ALT | 140.67 ± 629.12 | 58.95 ± 55.11 | 0.546 |
| WBC | 8.37 ± 3.48 | 9.54 ± 4.62 | 0.213 |
| Hg | 12.23 ± 1.85 | 11.49 ± 2.02 | 0.117 |
| PLT | 170.40 ± 94.30 | 176.55 ± 165.69 | 0.831 |
| Creatinine | 1.28 ± 0.71 | 1.44 ± 0.67 | 0.373 |

PT, prothrombin time; aPTT, activated partial thromboplastin time; FDP, fibrinogen degradation product; AST, aspartate aminotransferase; ALT, alanine aminotransferase; WBC, white blood cell; Hg, hemoglobin; PLT, platelet.

**Figure 1.**
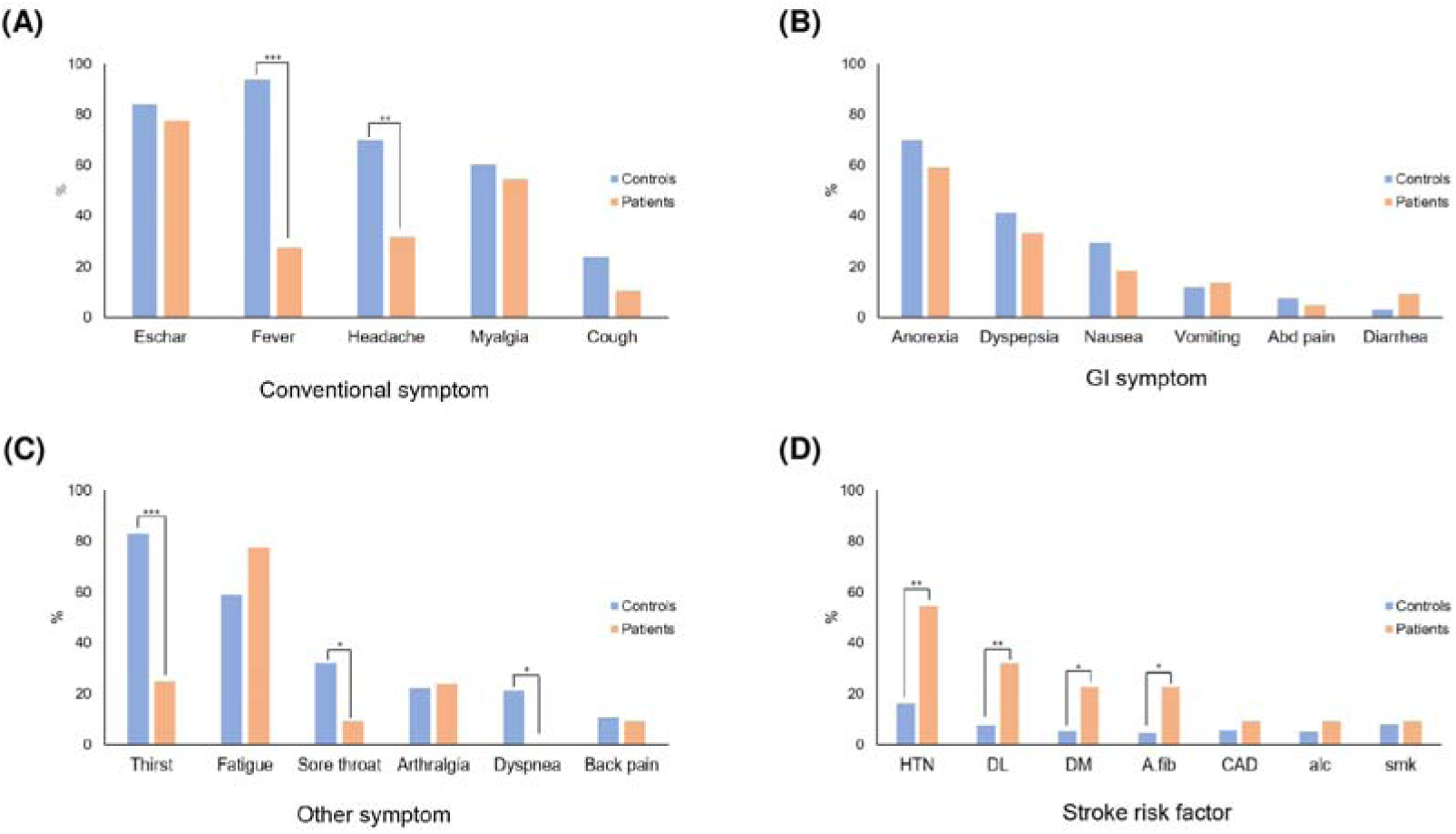
Comparison of symptoms and stroke risk factors between controls (tsutsugamushi without stroke) and patients (tsutsugamushi with stroke). Conventional symptoms of patients having tsutsugamushi with and without acute ischemic stroke (A). Gastrointestinal symptoms of patients having tsutsugamushi with and without acute ischemic stroke (B). Other symptoms of patients having tsutsugamushi with and without acute ischemic stroke (C). Stroke risk factors in controls and patients (D). A statistically significance is expressed as *, ** or ***. *, p<0.05; **, p<0.01; ***, p<0.001.

The present study performed multivariate analysis to identify factors associated with developing AIS in patients with tsutsugamushi (Table 4). Hypertension, diabetes mellitus, atrial fibrillation, and dyslipidemia, known risk factors for stroke, did not show significant results. However, higher D-dimer level at hospitalization due to tsutsugamushi was found to affect the development of AIS in tsutsugamushi patients (adjusted odds ratio [OR], 1.58; 95% confidence interval [CI], 1.06–2.34; p=0.023).

**Table 4.** Multivariate analysis of factors associated with tsutsugamushi with acute ischemic stroke

|  | **Crude OR (95% CI)** | ***P value*** | **Adjusted OR (95% CI)** | ***P value*** |
| --- | --- | --- | --- | --- |
| **Hypertension** | 6.27 (2.08–18.85) | *0.001* | 2.64 (0.24–29.65) | *0.432* |
| **DM** | 5.29 (1.15–24.49) | *0.033* | 5.86 (0.53–64.50) | *0.149* |
| **A.fib** | 5.29 (1.14–24.48) | *0.033* | 4.55 (0.61–33.75) | *0.139* |
| **Dyslipidemia** | 4.48 (1.24–16.21) | *0.022* | 0.95 (0.05–17.94) | *0.971* |
| **D-dimer** | 1.74 (1.22–2.47) | *0.002* | 1.58 (1.06–2.34) | *0.023* |

DM, diabetes mellitus; A.fib, atrial fibrillation; OR, odds ratio; CI, confidence interval.

## Discussion

This retrospective study aimed to determine the effects of tsutsugamushi infection on AIS onset. The co-occurrence of tsutsugamushi and AIS is rare; only a few cases have been reported till date.^12, 13^ The present study collected the data of 22 tsutsugamushi patients with AIS from three hub university hospitals over 15 years, compared them with the data of 66 tsutsugamushi patients without AIS (control), and identified the relationship between tsutsugamushi infection and AIS development.

Four of the 22 patients who developed AIS after the tsutsugamushi infection developed AIS >10 days after the onset of symptoms (Table 1). AIS diagnosis was delayed for Patient 1 because although the patient had syncope, an ischemic stroke symptom, 2 days before the diagnosis of tsutsugamushi, it was before hospitalization. The diagnosis was delayed in Patients 10 and 11 because they were not hospitalized in the neurology department, which delayed their neurological evaluation. Patient 14 developed AIS shortly after tsutsugamushi treatment and was discharged from the hospital, resulting in a late diagnosis. AIS was diagnosed in 18 patients after tsutsugamushi infection for <10 days (Table 1).

According to a recent multi-institute registry study in South Korea, in patients with stroke due to “other determined etiology” such as hypercoagulable state, the prevalence of stroke risk factors was 47.5, 23.8, 20.6, 3.2, and 21.2% for hypertension, dyslipidemia, diabetes, atrial fibrillation, and current smoker, respectively.^14^ The results of our study showed that the prevalence of risk factors was 54.5, 31.8, 22.7, 22.7, 9.1, and 9.1% for hypertension, dyslipidemia, diabetes mellitus, atrial fibrillation, coronary artery disease, and current smoking, respectively, which was generally higher than those of stroke patients due to “other determined etiology” (Table 3). This difference was likely because this study’s tsutsugamushi patients with AIS had an infectious disease (tsutsugamushi). The patients in the present study might have had more underlying diseases than the general population because people with multiple concomitant diseases or poor immunity due to an underlying disease are more likely to have an infectious disease.

The incidence of fever was significantly lower in tsutsugamushi patients with AIS than in those without AIS at admission (tsutsugamushi patients without AIS vs. tsutsugamushi patients with AIS; 93.8 vs. 27.3%, respectively; p<0.001; Table 3), which was believed to be related to the sequence of symptoms resulting from tsutsugamushi infection. Fever, one of the most common symptoms of tsutsugamushi, appears relatively early after infection,^15^ whereas coagulopathy is a late manifestation of the illness.^12^ Considering the sequence of these symptoms and the fact that fever protects the body against infection,^16^ fever development early in the course of tsutsugamushi infection is thought to trigger the body’s defense mechanism against *O. tsutsugamushi* in the early stages of infection. However, we suspect that patients who did not develop fever early in the course of their infection did not adequately deal with the bacteria; instead, the disease progressed to a coagulation disorder, which is a late manifestation. This hypothesis is in agreement with findings from previous studies that reported intravascular coagulation was more common in tsutsugamushi patients with severe illnesses than those with less severe illnesses.^4^ In addition to fever, the incidence rate of headache, sore throat, dyspnea, and thirst was significantly lower in tsutsugamushi patients with AIS than in those without AIS (Table 3, Figure 1). This finding was expected because headache, sore throat, dyspnea, and thirst are secondary fever symptoms.

Regarding stroke risk factors, tsutsugamushi patients with AIS had significantly higher rates of hypertension, diabetes mellitus, atrial fibrillation, and dyslipidemia than those without AIS (Table 3 and Figure 1). These results suggest that tsutsugamushi infection is more likely to cause AIS in patients with stroke risk factors than in those without the risk factors. Furthermore, several previous studies have reported that common acute infections trigger ischemic stroke.^3, 17^ However, we suspect that the triggering action of these infections targets stroke risk factors. For example, suppose a patient with underlying atrial fibrillation is infected with tsutsugamushi. In that case, a thrombus is more likely to develop due to increased heart rate in response to the infection, thus increasing the likelihood of AIS development. This corroborates with findings from the previous studies that reported thrombus formation was the pathogenesis of atrial fibrillation to induce AIS.^18^ The heart rate increases during infection, and when the resting heart rate increases, the risk of incident venous thromboembolism and inflammatory and coagulation factors also increases.^19^ This means that tsutsugamushi infection acts as a trigger, further aiding the patient’s pre-existing risk factors to cause AIS. Our hypothesis needs to be tested in a larger, systematic study in the future.

The target patient group’s aPTT, D-dimer, and FDP levels were significantly higher than those in the control group (p=0.009, 0.005, and 0.009, respectively). Elevated aPTT, D-dimer, and FDP levels were associated with coagulation disorders. Therefore, the results suggested that the mechanism of tsutsugamushi infection to induce AIS was related to coagulation diseases, which agreed with the results of previous study.^4^ Furthermore, from multivariate logistic analysis, we identified D-dimer elevation as a factor influencing the development of AIS after tsutsugamushi infection (adjusted OR, 1.58; 95% CI, 1.06–2.34; p=0.023). Dardiotis et al.^20^ reported that the secretion of inflammatory cytokines by cancer triggered an inflammatory response, which created a hypercoagulable state, resulting in systemic and cerebral arterial or venous thrombosis that could induce ischemic stroke. In addition, D-dimer elevation has been observed in patients with such cancer-related strokes, and brain images showed infarction of multifocal lesions.^20^ Moreover, Jackson et al.^21^ reported that inflammation and hemostatic response were activated in response to infection. Based on the results of these previous studies and the present study, we predicted that tsutsugamushi patients with a higher D-dimer level were more likely to develop AIS than those with a lower D-dimer level at the time of admission and progress to an embolic stroke due to their hypercoagulable state. After excluding three patients without imaging information, this study examined D-dimer levels in 19 of 22 AIS patients with tsutsugamushi infection and found that the D-dimer level of 16 patients exceeded the threshold of 0.5 mg/L (Table 2). Among the 16 patients, 13 showed an embolic pattern on brain MRI, which supported our hypothesis (Table 2). However, we could not conclude whether higher D-dimer levels induced embolic stroke. Because atrial fibrillation, another risk factor, can cause a cardioembolic stroke by abnormally promoting thromboembolism,^22^ the five patients with atrial fibrillation, elevated D-dimer level (≥0.5 mg/dL), and embolic pattern lesions on DWI could have developed cardioembolic stroke due to atrial fibrillation. However, the eight patients who showed embolic-pattern lesions with D-dimer elevation but without atrial fibrillation could support our proposed hypothesis. Notably, previous study has reported that tsutsugamushi infection can elevate coagulation factors, such as D-dimer, and cause DIC to induce bleeding;^4^ however, its association with ischemic stroke has not been reported. In addition, some previous studies have hypothesized that tsutsugamushi infection-induced coagulopathy affects the onset of stroke,^12, 13^ but all these studies were case reports. We believe that the novelty of the present study lies in its evaluation of the association between tsutsugamushi infection and AIS by analyzing brain images and data.

This study had several limitations. First, we could not control for all confounding variables because it was a retrospective study. Therefore, this study could not regulate variables other than tsutsugamushi infection that could cause AIS. Future prospective cohort studies are needed to improve the reliability of our results. Second, because the data were collected at a tertiary hospital, many patients had severe diseases, which could have introduced selection bias. However, because most people in South Korea visit tertiary hospitals when they suspect a stroke, finding a co-occurrence of tsutsugamushi and AIS in small primary care clinics is difficult. Considering these points, we believe the results will be similar even after including patients from primary and secondary hospitals. Third, it is necessary to exclude the possible influence of atrial fibrillation history on the stroke pattern to explain the correlation between D-dimer level and embolic patterns. However, this study could not compare patients with and without atrial fibrillation. It is necessary to conduct a histological study to demonstrate that excessive thrombin production due to DIC, rather than atherosclerosis, causes an embolic stroke. However, there was a limitation in that histological samples could not be obtained from stroke patients. Finally, the sample size was small because the co-occurrence of tsutsugamushi and AIS is rare. The statistics could have been biased because of the small sample size. For the same reason, we could not perform a statistical analysis to determine whether the brain MRI embolic pattern of the target group was associated with D-dimer levels. However, collecting data from 22 patients was meaningful because no other study has collected data from as many tsutsugamushi patients with AIS.

## Conclusion

On brain MRI, AIS in tsutsugamushi patients tended to appear in an embolic rather than a non-embolic pattern. They occurred more easily in patients with many stroke risk factors. Moreover, these patients were less likely to have a fever and had higher D-dimer levels. We predict that D-dimer plays a crucial role in the pathophysiology of tsutsugamushi infection and increases the occurrence of AIS. In this study, we tested the hypothesis that tsutsugamushi infection increased the likelihood of developing AIS in individuals with stroke risk factors. We surmise that if a patient does not develop a fever early in the course of the tsutsugamushi infection, the patient may develop AIS after a few days. In conclusion, if a tsutsugamushi patient has an elevated D-dimer level or does not have a fever on admission, the possibility of AIS should be considered.

## Data Availability

Yes, I include a statement.

## Acknowledgments

None.

